# Importance of Social Distancing: Modeling the spread of 2019-nCoV using Susceptible-Infected-Quarantined-Recovered-t model

**DOI:** 10.1101/2020.04.17.20069245

**Authors:** Nipun Aggarwal

**Author notes:** Email address (Nipun Aggarwal).

## Abstract

**Background:** The Novel Coronavirus that originated in Wuhan, Hubei, China, has raised global concerns and has been declared a pandemic. The infection shows the primary symptoms of pneumonia and has an incubation period, with the majority of people showing symptoms within 14 days. Online Social Networks are the closest simulations of real-world networks and have similar topology characteristics. This article simulates the spread and control of the nCoV-19 using the SIQR-t model to highlight the importance of self-quarantine and exercise of proper health care as a method to prevent the spread of the virus.

**Method:** The article uses the Susceptible-Infected-Quarantined-Recovered model with modification, introducing 14 different Infected states depending on the number of days the host has been carrying the infection. We simulate the spread of 2019-nCoV on human interaction similar graph taken from Online Social Network Epinions, of about 75000 nodes, similar to a small town or settlement. The infection rates depend on the sanitation and cleanliness these people exercise.

**Results:** When people practice self-quarantine and hygiene, aided by the governmental efforts of testing and quarantine, the cumulative number of affected people fall drastically. The decrease is apparent in time-based simulations of the spread received from the study.

**Conclusion:** The 2019-nCoV is a highly infectious zoonotic virus. It has spread like a pandemic, and governments across the world have launched quarantines. The results of the SIQR-t model indicate that hygiene and social-distancing can reduce its impact and sharply decrease the infection scale. Individual efforts are key to the control.

## 1. Introduction

The Novel Coronavirus that originated in Wuhan City, Hubei Province of China, has been declared a global pandemic [1, 2]. It is majorly spread through persistence on inanimate surfaces and is similar to outbreaks of SARS and MERS, originating through bats [3, 4]. Studies on the nature of the virus have suggested different incubation periods of the virus, and reports have suggested a median incubation period of 5-6 days and a very high symptom probability period of 14 days [5]. The virus is transmitted through carriers even before their symptoms show [6]. The article suggests the simulation on Online Social Networks that resemble human interactions, using time-variant infection rates that accommodate the dormancy and incubation period of the novel virus, to simulate the spread of the virus among people. This research aims to provide a mathematical model for the spread and control of the virus, considering the control measures of quarantine and social distancing. The results shall highlight the importance of early-detection, self-quarantine and hygiene practices.

## 2. Traditional Model

The methods used in this paper involve the simulation on Epinions social network as the topology of spread, using the general consumer review website with the same name. Members of the site can decide whether to trust each other, thus simulating the interactions between people in a total population of 75879 with about 508837 connections in these people representing potential transfer among these people [7]. The traditional model used to study diseases is usually the Susceptible-Infected-Recovered-model, which is a popular model to simulate outbreaks, including the 2019-nCoV [8]. This model may be extended to include a quarantine state to give the Susceptible-Infected-Quarantine-Recovered model [9]. This paper uses a modified version of this model, considering that the infection and quarantine (on symptom) rate varies based on the number of days, the host is carrying the virus [10]. Based on the findings regarding the spread of the virus, 14 different infection and quarantine rates are hyper-parameters that are based on the hygiene and social-distancing practices assumed for these hosts. The paper’s contributions are as follows:

- Mathematical model for a time-varying virus infection rate
- Importance of self-quarantine and early detection of symptoms
- Potential model for application to predicting the spread and control of 2019-nCoV

The rest of the paper is organized into sections. Section 3 proposes the mathematical model of the SIQR-t model. The application to a real-life network is simulated in Section 4, and the importance of self-quarantine is highlighted. Section 5 concludes the applicability of the model to the novel coronavirus.

## 3. SIQR-t Model

The proposed SIQR-t model has the states as given in Fig. 1

The states of the model and the transitions are given as:

**Figure 1:**
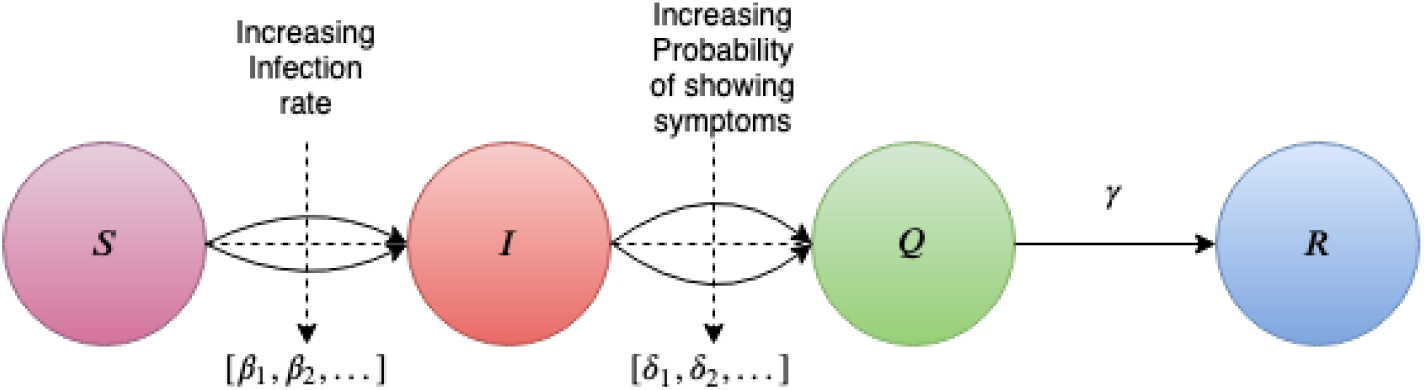
*SIQR* − *t* Model

- All people are susceptible to the disease and start as Susceptible Nodes. Some of the Nodes have acquired the disease and are active spreaders, Infected Nodes, at the beginning of the simulation.
- The infected nodes infect their neighbors at rate *β*_*i*_, where *i* denotes the days passed since their infection.
- The infected nodes begin to show symptoms at rate *δ*_*i*_, where *i* denotes the days passed since their infection.
- After reaching the quarantine, the host tends to recover at the rate *γ* slowly and stops infecting people due to limited contact with the outside world.

### 3.1. System of differential equations

The population is divided into the 4 states Susceptible *S*, Quarantined *Q*, Infected *I* and Recovered *R* and at any time *t* their sum is 1 as in Eq. 1.

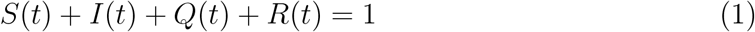

As evident from Fig. 1, some infected nodes may become quarantined at varying rates *δ*_*i*_ as per the Eq. No. 2. Quarantined nodes are those hosts who are no longer interacting with people. The node in the quarantined(*Q*) state tries to heal itself and practices heavy social isolation.

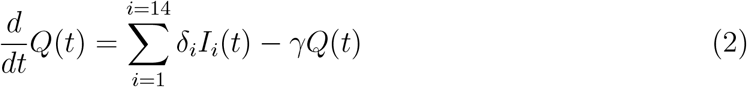

Infected nodes increase at multiple different rates, either moving to the next day, becoming newly infected or becoming quarantined (edge between *I* to *Q* in the Fig. 1). The state for *I*_1_ in Eq. 3 is given separately and a general formula for others is provided in Eq. 4.

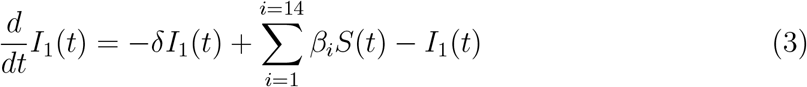

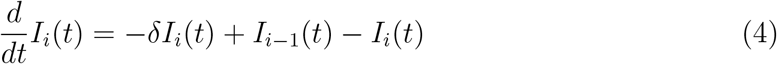

The transition into recovered nodes, Eq 5, arises from the conversion of quarantined nodes to recovered nodes with the rate *γ*

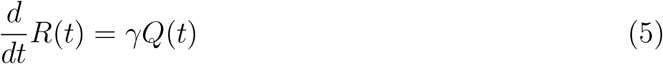

Initially, all the nodes are assumed susceptible except few seed nodes and slowly, they decay into infected state Eq. 6.

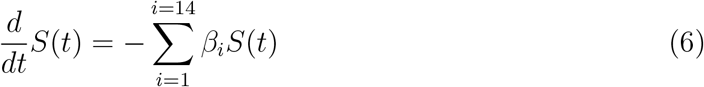

## 4. Simulation and Results

The model was coded and simulated using NetworkX [11], and the code is publicly available at Github repository [12]. It is simulated on the Epinion network [7]. This network is ideal for representing a small settlement or town with an average diameter of 5 nodes.

As evident in Fig. 2(a), people are likely to get infected and are quarantined after confirmation of their symptoms. The spread is high, and the quarantine starts to shoot up only after about 35,000 of the total population is already actively infected.

**Figure 2:**
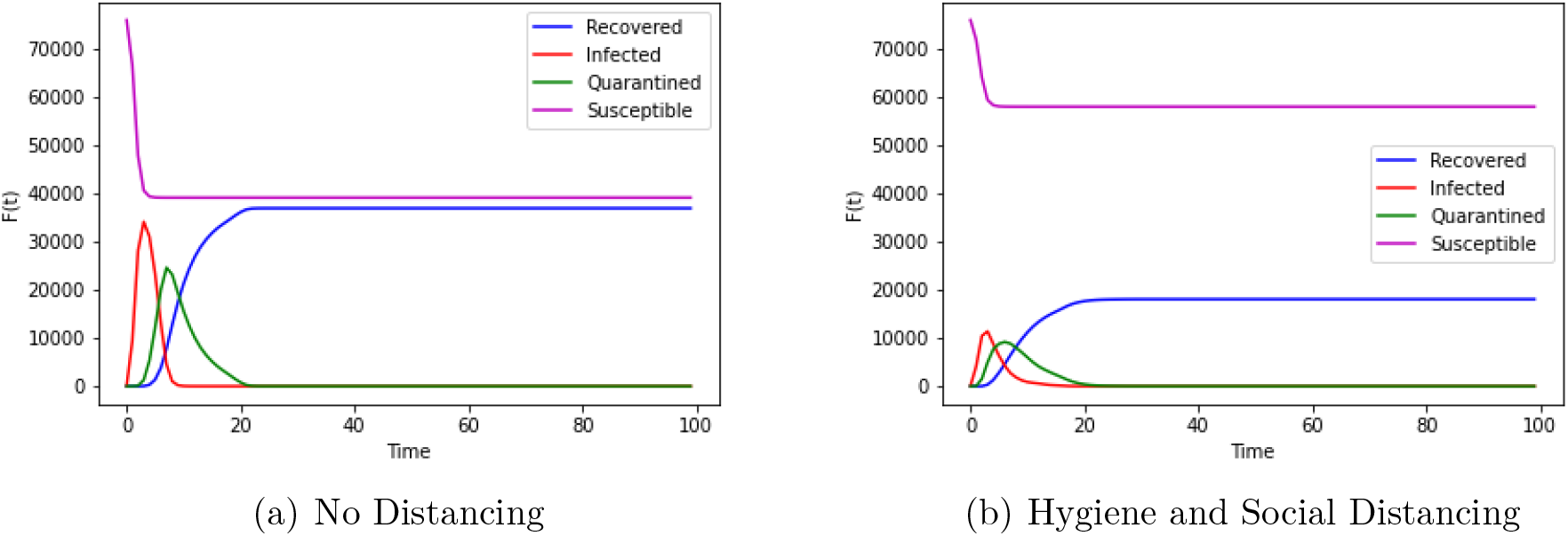
All nodes are initially susceptible and an influential node is chosen as the spreader.

If people practice minimal social interaction, regardless of their infection and take extra precautions on their infection, the expected peak of infection drops rapidly, and the overall count of nodes affected by the virus is decreased manifold, as evident in Fig. 2(b).

## 5. Conclusion

The SIQR-t model is a set of differential equations aimed at simulating the spread of 2019-nCoV, creating awareness for sanitation practices, social-distancing and quarantine to prevent the spread of the virus. It suggests the usage of varying rates of infection slowly increasing as the infection progresses. The probability that the host quarantines itself increase as each day passes because the dormant symptoms become evident. As suggested by the findings of the research, people can use social distancing and hygiene practices to reduce the negative effects of the pandemic radically.

## Data Availability

All the data used in the paper is publicly available and properly cited in the manuscript

https://snap.stanford.edu/data/

https://github.com/DeveloperAlfa/nCoV-19

